# A Systematic Review and Meta-Analysis of the Mental Health Symptoms during the Covid-19 Pandemic in Southeast Asia

**DOI:** 10.1101/2021.06.03.21258001

**Authors:** Sofia Pappa, Jiyao Chen, Joshua Barnet, Anabel Chang, Rebecca Kechen Dong, Wen Xu, Allen Yin, Bryan Z. Chen, Andrew Delios, Richard Z. Chen, Saylor Miller, Xue Wan, Stephen X. Zhang

## Abstract

**Aims:** The Covid-19 pandemic has had a substantial impact on the mental health of the general public and high-risk groups worldwide. Due to its proximity and close links to China, Southeast Asia was one of the first regions to be affected by the outbreak. The aim of this systematic review was to evaluate the prevalence of anxiety, depression and insomnia in the general adult population and healthcare workers (HCWs) in Southeast Asia during the course of the first year of the pandemic.

**Methods:** Several literature databases were systemically searched for articles published up to February 2021 and two reviewers independently evaluated all relevant studies using pre-determined criteria. The prevalence rates of mental health symptoms were calculated using a random-effect meta-analysis model.

**Results:** In total, 32 samples from 25 studies with 20,352 participants were included. Anxiety was assessed in all 25 studies and depression in 15 studies with pooled prevalence rates of 22% and 16% respectively. Only two studies assessed insomnia, which was estimated at 19%. The prevalence of anxiety and depression was similar amongst frontline HCWs (18%), general HCWs (17%), and students (20%) whilst being noticeably higher in the general population (27%).

**Conclusions:** This is the first systematic review to investigate the mental health impact of the Covid-19 pandemic in Southeast Asia. A considerable proportion of the general population and HCWs reported mild to moderate symptoms of anxiety and depression; the pooled prevalence rater, however, remain significantly lower than those reported in other areas such as China and Europe.

## 1. Introduction

As of May 2021, 155.3 million cases of Covid-19 had been confirmed, resulting in 3.2 million deaths worldwide [1]. Southeast Asia, comprising 11 countries and over 670 million people), was the first region outside of the initial outbreak in China to report Covid-19 cases on January 13^th^ 2020 in Thailand [2], and deaths on February 2^nd^ 2020 in The Philippines [3].

Southeast Asian states share extensive ties with China. For instance, the annual travellers between Singapore (a Southeast Asian country of 3.5 million citizens) and Wuhan (the epicentre of the Covid-19 outbreak) number around 3.4 million [4]. Furthermore, many Southeast Asian countries are developing countries with high population density and potentially lacking in resources, healthcare personnel, or facing challenges to enforce social distancing and lockdowns [5-7]. Nonetheless, the region has had several recent experiences with high-profile epidemics, such as SARS in 2003 and H1N1 in 2009, which may have led to better public and medical preparedness and pandemic response in Southeast Asia [8].

Indeed, previous reports have demonstrated high rates of adverse mental health symptoms in the general population and in vulnerable groups during past infectious disease outbreaks [9]. Furthermore, a number of rapid reviews and recent meta-analyses have established the pooled prevalence of mental health disorders during the Covid-19 crisis in China and other areas [10-15]. However, the region of Southeast Asia, despite its vast population, proximity to China and recent experiences with prior epidemics, has not received a meta-analysis on the mental health symptoms during the Covid-19 pandemic. In order to fill the gap in the evidence, this systematic review aims to evaluate the pooled prevalence rates of anxiety, depression and insomnia in the general public, healthcare workers and students during the first year of the pandemic in Southeast Asia.

The one-year scope of the systematic review and meta-analysis allowed for a broad evidence-based assessment of all the available data, in order to produce a set of pooled prevalence of the key mental health symptoms studied to date, and helped to address the effect of sample size bias and the heterogeneity of results between studies [16] (Pappa et al. 2021a). Furthermore, this systematic review covers the mental health impact of the general adult population, as well as healthcare workers (HCWs) and students, who have been reportedly at greater risk of experiencing mental health difficulties during the Covid-19 pandemic [17,18]. The findings of this study can contribute to the existing body of research on the subject to facilitate comparisons with other regions and inform evidence-based practice and epidemic planning of mental health needs.

## 2. Methods and Materials

The review protocol is registered with the International Prospective Register of Systematic Reviews (PROSPERO: CRD42020224458) and the systematic search was conducted according to the Preferred Reporting Items for Systematic Reviews and Meta-Analyses (PRISMA) statement 2019.

### 2.1 Data Sources and Search Strategy

This work is part of an overarching project of large-scale meta-analyses of the psychological impact of the Covid-19 pandemic on the general and high-risk populations across the globe. We searched the following databases for studies that met the inclusion criteria: *PubMed, Embase, PsycINF*O, and *Web of Science* from Feb 1^st^, 2020 to Feb 6^th^, 2021. Preprints published at *medRxiv* were also included in the search. To identify articles based on the requirements, we searched specific titles and abstracts using the search terms in Table S1 with Boolean operators.

### 2.3 Selection Criteria

The search included empirical studies that reported on the prevalence of anxiety, depression, or insomnia among frontline HCWs, general HCWS, general adult populations, or adult (university) students in Southeast Asia. In addition, reports had to employ validated psychometric measures and outcomes had to be reported in English.

We did not include studies that reported on populations of children, adolescents, or adult subpopulations (e.g., pregnant women). Non-original research or studies which were reviews, meta-analyses, qualitative and case studies, interviews, news reports, interventional studies, or studies that did not use validated instruments or validated cut off scores to quantify prevalence rates were also excluded.

A researcher (WX) contacted the authors of papers that missed important information in several instances: 1) if they surveyed a population that included both targeted and excluded populations in a way that we could not identify the prevalence rate for our desired population; 2) if the paper included primary data meeting our inclusion criteria, but did not report the prevalence; 3) if the paper reported the overall prevalence without specifying whether it is mild above or moderate above: or 4) if the paper was missing or unclear about critical information such as respondent rate, data collection time, or female proportion rate.

### 2.3 Data Screening

Article information from various databases was initially extracted into Endnotes to remove duplicates and then imported into Rayyan. Two researchers independently (BZC & AD) screened the titles and abstracts of all papers against the inclusion and exclusion criteria. Potential conflicts were resolved by a third researcher (RKD).

### 2.4 Data Extraction

A well-developed coding protocol and coding book were used based on previous studies [19]. All included articles from the screening were assigned to three pairs of researchers (WX & AY, BZC & AD, RZC & SM) who thoroughly examined and extracted important data into a coding book. Relevant information including author, title, country, starting and ending dates of data collection, study design, population, sample size, respondent rate, female proportion rate, age range and mean, outcome, outcome level, instruments, cut-off scores, and prevalence were coded using a standard coding procedure. Comments and reasons for contacting authors and/or excluding papers were also recorded.

After both coders had independently coded their articles, they would crosscheck their information. In the event of disparities and in the absence of a consensus between the two, a third coder would settle the disagreement. The third coder (AZ) did also double-check important data including the population, sample size, mental health outcomes, outcome levels, instruments, and prevalence. Studies with unusual prevalence, cut off scores, and numbers were afterwards also checked in the sensitivity analysis.

### 2.5 Assessment of Risk of Bias

The Mixed Methods Appraisal Tool (MMAT) [20] of seven questions was used to assess the quality of research papers included in the meta-analyses. The tool consists of seven questions and quality scores range from 0 to 7. Studies with a score of above 6 were considered high quality, between 5 and 6 were classed as medium, and with a score of below 5 were considered of low quality. Questions were individually coded by pairs of coders and any discrepancies resolved by a third coder (RKD).

### 2.6 Data analysis

Version 16.1 of Stata was utilized and a random effect meta-analysis conducted to extract the pooled prevalence from multiple studies using meta-prop. We used the *I*^*2*^ statistics to examine the heterogeneity of prevalence among studies and the heterogeneity was classed as high when *I*^*2*^ was higher than 75% [21].

## 3. Results

### 3.1 Study Selection

Figure 1 illustrates the PRISMA flow chart detailing our search and study retrieval process and findings. In total, 6949 records were screened for their title, abstract and keywords. After removal of duplicates, we reviewed 168 articles by their full texts against our eligibility criteria. Finally, we analyzed data from 25 studies satisfying the inclusion criteria.

**Figure 1.**
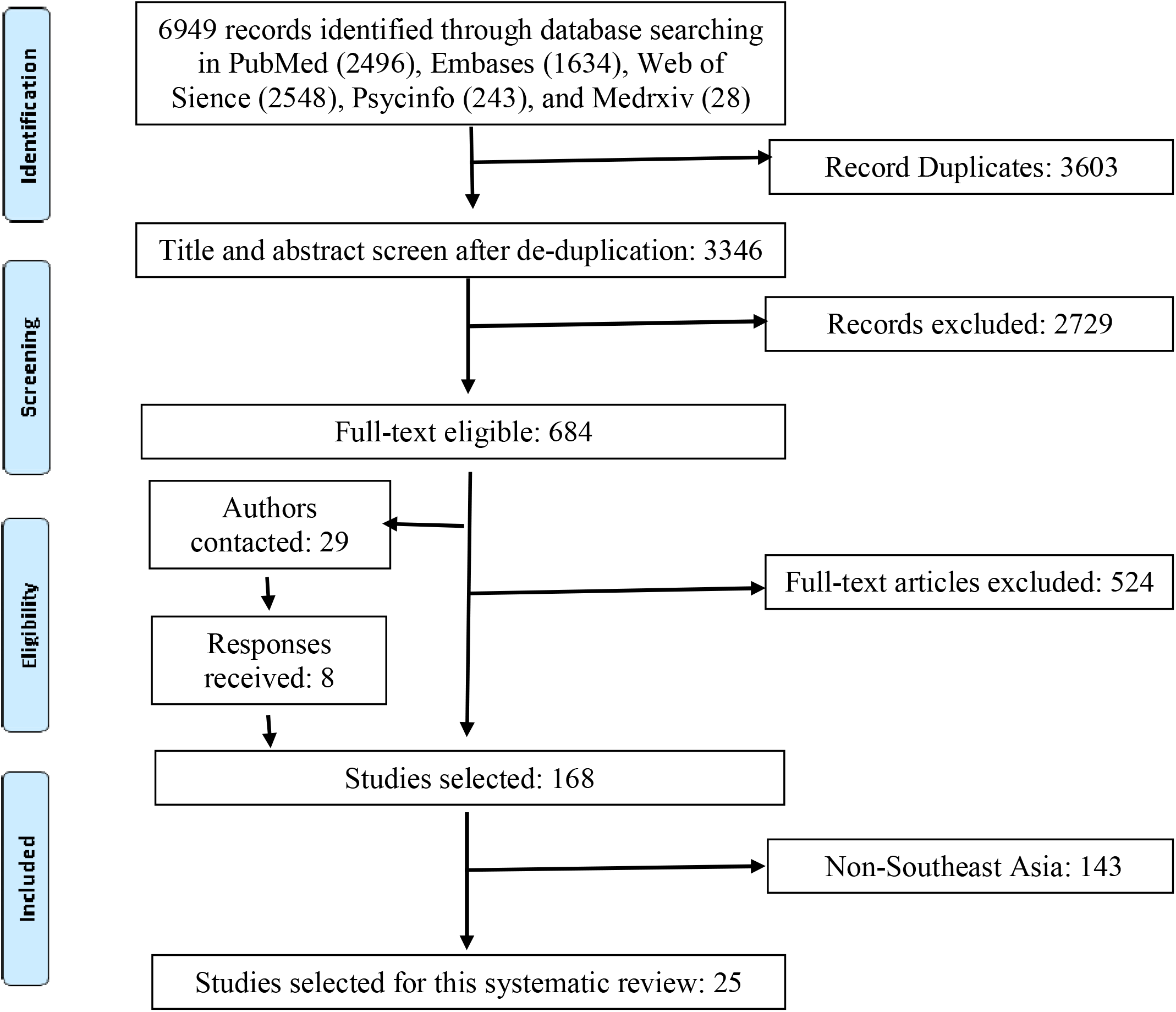
PRISMA flow diagram.

### 3.2 Study Characteristics

In total, 25 studies including 32 different samples and 20,352 participants from Southeast Asia were incorporated in this meta-analysis [22-46]. Of these, 7 studies (28.1%) were of general populations [25,28,30,34,43,44,46], 10 studies included general HCWs (43.8%) [19,22,24,29,31-33,37,40,45], 5 studies included frontline HCWs (18.8%) [27,37-39,41], and only 3 studies (9.4%) referred to adult students [26,36,47]. Most studies were cross-sectional (96.0%) apart from one that was a longitudinal cohort study (4.0%) (Wong et al. 2021). The sample size across all 32 samples varied from 22 to 4004 and a median number of 294 and the proportion of female participants ranged from 47.6% to 88.1% with a median value of 69.5%. The participation rates were between 20.0% to 98.0% and a median of 70.3%. All studies had been published.

### 3.3 The pooled prevalence of anxiety, depression, and insomnia

All 32 samples with a total of 20,352 participants from the 25 studies reported on the prevalence of anxiety symptoms. Several validated assessment tools were used, including most commonly the Depression, Anxiety and Stress Scale - 21 Items (DASS-21) (37.5%), followed by the Generalized Anxiety Symptoms 7-items scale (GAD-7) (25.0%), the Hospital Anxiety and Depression Scale (HADS) (16.7%), and six other instruments (each 4.2%). Different studies used different cut-off values to determine the overall prevalence as well as the severity of anxiety. In the random-effects model, the pooled prevalence rate was 22% (95% CI: 19% - 27%, I2 = 99.9%) (Figure 2A).

**Figure 2A.**
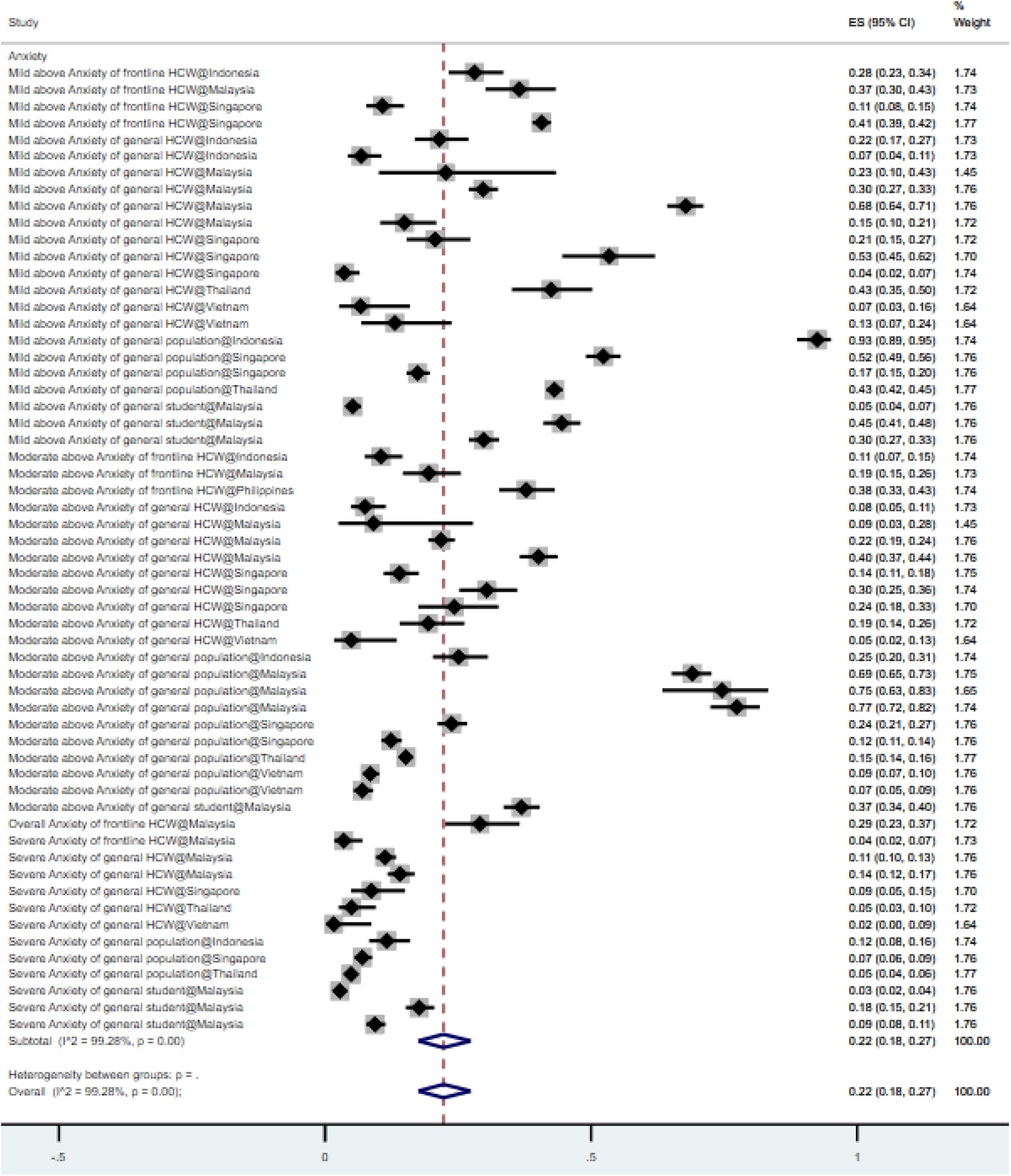
Forest plot of the prevalence of anxiety. Figure legend: The square markets indicate the prevalence of anxiety at the different level for different population. The size of the marker correlates to the inverse variance of the effect estimate and indicates the weight of the study. The diamond data market indicates the pooled prevalence.

A total of 20 samples and 13,960 respondents deriving from 15 studies [23,24,28-32,35,37,38,40,41,43,45,47] reported in the presence and severity of depression. A variety of rating scales were used such as DASS-21 (60.0%), HADS (20%), and three other instruments (each 6.7%). In the random-effects model, the pooled prevalence of depression was 16% (95% CI: 12% - 20%, I2 = 99.8%) (Figure 2B).

**Figure 2B.**
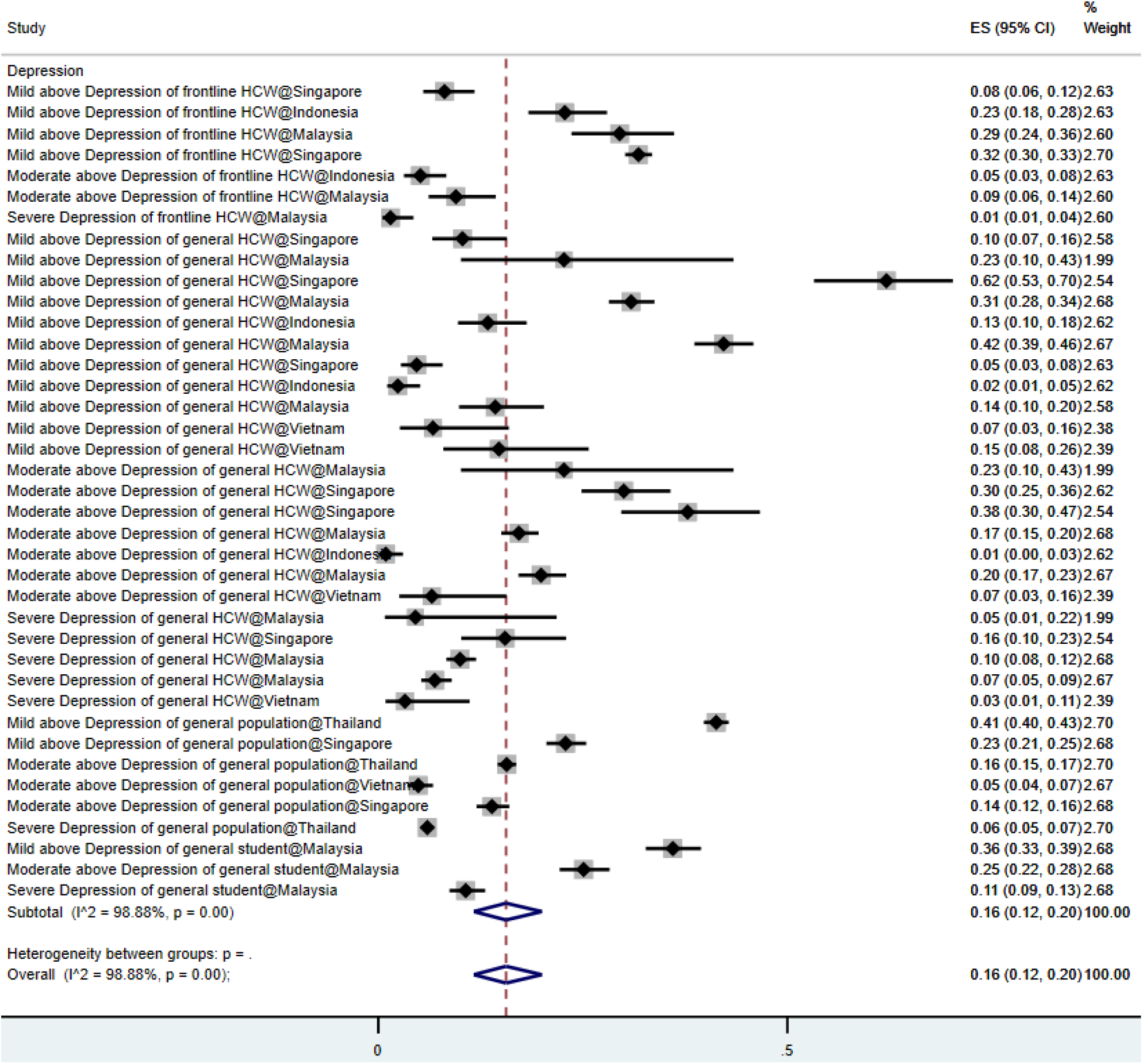
Forest plot of the prevalence of depression. Figure legend: The square markets indicate the prevalence of anxiety at the different level for different population. The size of the marker correlates to the inverse variance of the effect estimate and indicates the weight of the study. The diamond data market indicates the pooled prevalence.

The overall prevalence of mental disorder symptoms in frontline HCWs, general HCWs, students and the general population in Southeast Asia are 18%, 17%, 20%, and 27% respectively. The overall prevalence rates of mental disorder symptoms that surpassed the cut off values of mild, moderate, and severe symptoms were 26%, 21%, and 7%, respectively.

Only two samples from two studies [29,43] included the prevalence of insomnia with a pooled rate of 19%, hence insomnia is not included in the different sub-analysis presented here.

### 3.5 Quality of articles

According to the Mixed Methods Appraisal Tool (MMAT) [20], seven studies (28.0%) are found to be of high quality and the remaining 18 studies (72.0%) fall into medium quality (Table 1). The subgroup analysis indicated that studies with higher quality reported lower prevalence of mental health problems in Southeast Asia (p=0.00) (Table 2).

**Table 1.** Study Characteristics on mental health symptoms in COVID-19 epidemic in Southeast Asia.

| Characteristics | Total number of studies/samples* | Percent (%) | Level of analysis |
| --- | --- | --- | --- |
| *Overall* | 25/32 | 100 |  |
| *Design* |  |  | Study |
| Cohort | 1 | 4.0 |  |
| Cross-sectional | 24 | 96.0 |  |
| *Publication status* |  |  | Study |
| Preprint | 0 | 0.0 |  |
| Published | 25 | 100.0 |  |
| *Quality* |  |  | Study |
| >6 | 7 | 28.0 |  |
| Between 5 and 6 | 18 | 72.0 |  |
| <5 | 0 |  |  |
| *Population* |  |  | Study |
| Frontline HCW | 5 | 20.0 |  |
| General HCW | 10 | 40.0 |  |
| General population | 7 | 28.0 |  |
| Student | 3 | 12.0 |  |
| *Outcome#* |  |  | Prevalence |
| Anxiety | 58 | 59.8 |  |
| Depression | 39 | 40.2 |  |
| *Severity#* |  |  | Prevalence |
| Above mild | 41 | 42.3 |  |
| Above moderate | 35 | 36.1 |  |
| Severe | 20 | 20.6 |  |
| Overall | 1 | 1.0 |  |
| *Country* |  |  | Sample |
| Indonesia | 4 | 12.5 |  |
| Malaysia | 12 | 37.5 |  |
| Philippines | 1 | 3.1 |  |
| Singapore | 9 | 28.1 |  |
| Thailand | 2 | 6.3 |  |
| Vietnam | 4 | 12.5 |  |
|  | Median (mean) | Range |  |
| *Sample size* | 294 (636) | 22 - 4004 | Sample |
| *Response rate* | 77.7% (70.9%) | 17.4% - 100% | Sample |
| *Female portion* | 69.5% (68.3%) | 47.6% - 88.1% | Sample |

\* One study may include multiple independent samples. For example, Chew et al (2020) studies the prevalence in the general population of Singapore, Indonesia, Malaysia, and Vietnam

**Table 2.** The pooled prevalence rates of mental health symptoms by subgroups of population, outcome, and severity.

| First-level subgroup | Second-level subgroup | Prevalence (%) | 95% CI | I<sup>2</sup> (%) | P value |
| --- | --- | --- | --- | --- | --- |
| *Overall* |  | 20% | 16% - 23% | 99.2% | 0.00 |
| *Population* | Frontline HCW | 18% | 12% - 25% | 98.3% | 0.00 |
|  | General HCW | 17% | 13% - 21% | 97.9% | 0.00 |
|  | General population | 27% | 19% - 35% | 99.7% | 0.00 |
|  | Student | 20% | 11% - 30% | 99.2% | 0.00 |
| *Outcome#* | Anxiety | 22% | 18% - 27% | 99.3% | 0.00 |
|  | Depression | 16% | 12% - 20% | 99.9% | 0.00 |
|  | Insomnia | 19% | 1% - 23% | 99.3 | 0.00 |
| *Severity#* | Mild | 26% | 21% - 31% | 98.9% | 0.00 |
|  | Moderate | 21% | 16% - 26% | 98.6% | 0.00 |
|  | Severe | 7% | 6% - 9% | 93.7% | 0.00 |
| *Study Quality* | High quality | 16% | 11% - 21% | 98.5% | 0.00 |
|  | Medium quality | 21% | 17% - 25% | 99.3% | 0.00 |
| *Subregion* | Continental | 20% | 16% - 23% | 99.2% | 0.00 |
|  | Malay Archipelago | 18% | 8% - 31% | 98.9% | 0.00 |

Note: CI = Confidence Interval; I<sup>2</sup> statistic indicates the heterogeneity. Some I<sup>2</sup> values in Table 2 and Table 3 are missing because the total sample is less than 3.

### 3.6 Sensitivity Analysis

The use of conventional funnel plots to assess biases in meta-analyses have been found to be inaccurate for meta-analyses of proportion studies [48], for which the Doi plot and the Luis Furuya–Kanamori (LFK) index denote a better approach for graphically representing publication bias – where a symmetrical triangle implies the absence of publication bias, while an asymmetrical triangle indicates possible publication bias [49]. The Doi plot and LFK index have higher sensitivity and power to detect publication bias than the funnel plot and Egger’s regression [50]. The LFK index provides a quantitative measure to assess the asymmetry - a score within ±1 indicates ‘no asymmetry’, exceeds ±1 but is within ±2 indicates ‘minor asymmetry’ and exceeds ±2 indicates ‘major asymmetry’. Figure 3 depicts the Doi plot and a Luis Furuya–Kanamori (LFK) index of -1.75, indicating ‘minor asymmetry’ and the presence of minor publication bias. Moreover, specifically, we tested the impact of publication status and sample size and did not find significant influence.

**Figure 3.**
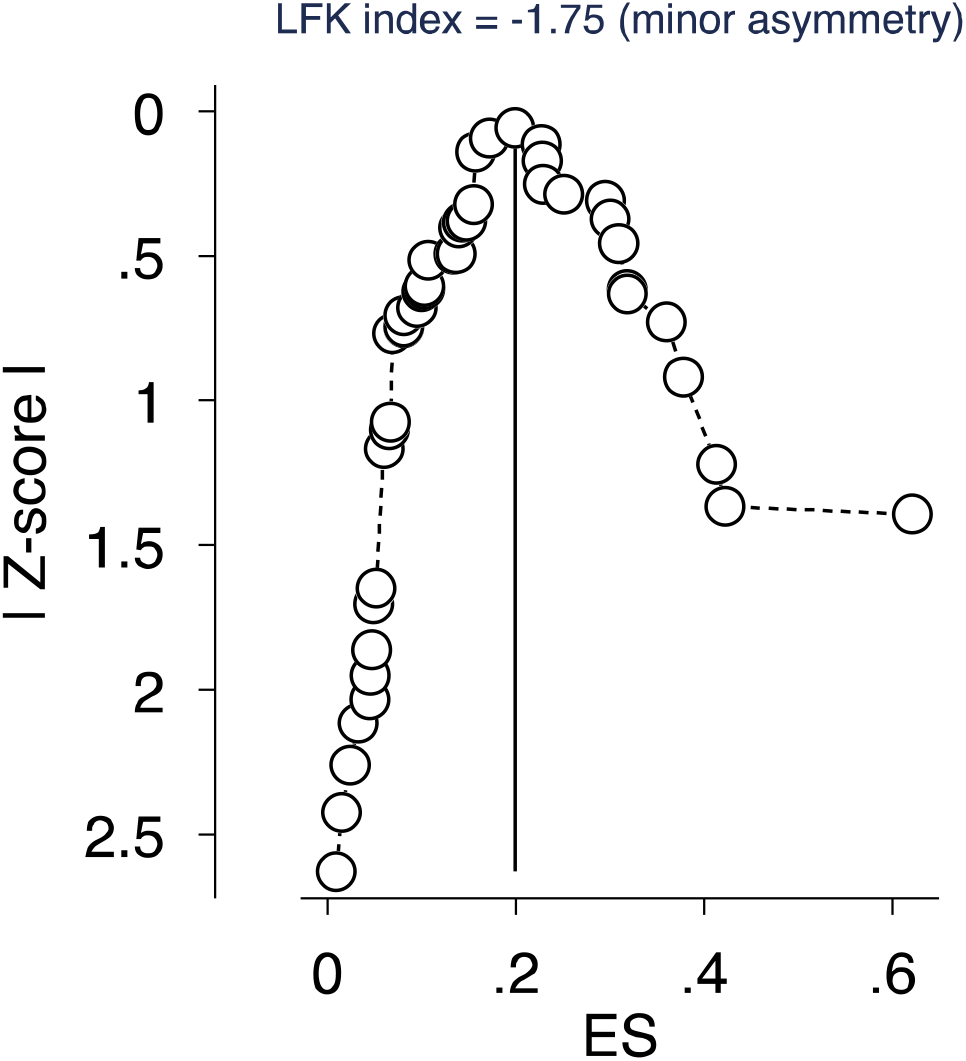
Depiction of publication bias in the baseline meta-analysis of proportion studies based on Doi plot and the Luis Furuya–Kanamori (LFK) index -a score that exceeds ±1 but is within ±2 indicates ‘minor asymmetry’

## 4. Discussion

### 4.1 Overview of Findings

This is the first systematic review with meta-analysis to assess the prevalence of mental health symptoms in the adult general and high-risk populations of Southeast Asia during the Covid-19 crisis. It included 32 samples from 25 studies for an aggregate of 20,352 adult participants in a year of the Covid-19 pandemic. Our findings showed that the overall prevalence of mental disorder symptoms was similar amongst frontline HCWs (18%), general HCWs (17%) and students (20%) whilst being noticeably higher in the general population (27%). Factors contributing to adverse psychological outcomes amongst the general population could include increased exposure to Covid-19 information from the media [25,30] urban living [25,44] and higher levels of perceived susceptibility to Covid-19 [46].

The pooled prevalence rates of anxiety, depression and insomnia were 22%, 16% and 19% respectively (Table 2). Anxiety was more prevalent in the general population than in HCWs and more frequent compared to depression in both groups (Table 3). Surprisingly, the overall level of moderate anxiety (21%) was not dissimilar to that of mild anxiety (26%). Regarding geographical distribution, there was a lower prevalence of depressive symptoms among the adults in the Malay Archipelago than those in continental Southeast Asia (7% vs 17%) despite comparable levels of anxiety (Table 3).

**Table 3.** Subgroup analyses of the prevalence of anxiety and depression.

| Groups | Subgroups | Anxiety | Depression |
| --- | --- | --- | --- |
| *# of studies* |  | 25 | 15 |
| *# of samples* |  | 32 | 20 |
| *# of prevalence* |  | 58 | 39 |
| *Total # participants* |  | 20352 | 13,960 |
| *Overall* |  | 22%, 95% CI: 19% - 27%, I<sup>2</sup>: 99.9% | 16%, 95% CI: 12% - 20%, I<sup>2</sup>: 99.9% |
| *Population* | Frontline HCW | 23%, 95% CI: 13% - 34%, I<sup>2</sup>: 98.1% | 14%, 95% CI: 5% - 25%, I<sup>2</sup>: 98.5% |
|  | General HCW | 18%, 95% CI: 12% - 80%, I<sup>2</sup>: 98.1% | 15%, 95% CI: 10% - 21%, I<sup>2</sup>: 97.5% |
|  | General population | 31%, 95% CI: 20% - 44%, I<sup>2</sup>: 99.7% | 16%, 95% CI: 6% - 29%, I<sup>2</sup>: 99.7% |
|  | Student | 18%, 95% CI: 8% - 32%, I<sup>2</sup>: 99.4% | 23%, 95% CI: 10% - 39% |
| *Severity* | Mild | 29%, 95% CI: 21% - 37%, I<sup>2</sup>: 99.1% | 21%, 95% CI: 16% - 28%, I<sup>2</sup>: 98.2% |
|  | Moderate | 25%, 95% CI: 18% - 33%, I<sup>2</sup>: 98.9% | 14%, 95% CI: 10% - 19%, I<sup>2</sup>: 96.6% |
|  | Severe | 8%, 95% CI: 5% - 11%, I<sup>2</sup>: 95.5% | 7%, 95% CI: 5% - 10%, I<sup>2</sup>: 87.9% |
| *Instrument* | DASS-21 | 17%, 95% CI: 12% - 24%, I<sup>2</sup>: 98.7% | 14%, 95% CI: 10% - 18%, I<sup>2</sup>: 97.7% |
|  | GAD-7 | 21%, 95% CI: 12% - 31%, I<sup>2</sup>: 99.6% | NA |
|  | HADS | 25%, 95% CI: 14% - 39%, I<sup>2</sup>: 97.8% | 18%, 95% CI: 7% - 33%, I<sup>2</sup>: 98.1% |
| *Region* | Continental | 22%, 95% CI: 17% - 27%, I<sup>2</sup>: 99.3% | 17%, 95% CI: 13% - 22%, I<sup>2</sup>: 98.9% |
|  | Malay Archipelago | 25%, 95% CI: 10% - 45%, I<sup>2</sup>: 99.1% | 7%, 95% CI: 2% - 16%, I<sup>2</sup>: 96.5% |

Note: CI = Confidence Interval; I<sup>2</sup> statistic indicates the heterogeneity.

### 4.2 Comparison of Results with Previous Studies

The prevalence rates of anxiety, depression and insomnia are overall lower in Southeast Asia than those reported in previous meta-analyses and studies from other areas or countries during the pandemic. They are considerably lower than, for example, the rates reported by the same study group covering the first year of the pandemic in Spain (34%, 36% and 52%) [51] and Africa (37%, 34% and 28%) [52] as well as by a separate meta-analysis from China (26%, 26%, and 30%) [14]. Likewise, the pooled prevalence of anxiety (22%) and depression (16%) in Southeast Asia was found to be consistently lower than the recorded scores of 33% and 32% for anxiety and 28% and 34% for depression in the meta-analysis by Luo et al. [53] from 17 countries (China, Singapore, India, Japan, Pakistan, Vietnam, Iran, Israel, Italy, Spain, Turkey, Denmark Greece, Argentina, Brazil, Chile and Mexico), and the meta-analysis by Salari et al. [54] from 10 countries (China, India, Japan, Iran, Iraq, Italy, Nepal, Nigeria, Spain, and UK), respectively. Furthermore, the pooled estimates from Southeast Asia are lower than the mental health outcomes previously reported among the general population and HCWs during and after the MERS and SARS epidemics where high rates of mood symptoms and post-traumatic stress disorder (PTSD) were observed [27,35,38,39,41].

The prevalence of psychological distress amongst students in Southeast Asia (20%), although deriving from a limited number of studies, compares favourably to that in Spain (50%) [51], a meta-analysis performed on studies from China, Iran, India, Brazil and the UAE (28% pooled prevalence of anxiety) [55] and a further meta-analysis from 31 countries performed by Deng et al. (anxiety 32%, depression 34%, insomnia 33%) [56].

The presence of mental health symptoms in HCWs in Southeast Asia follows a similar pattern compared with, for example, the first rapid systematic review and meta-analyses of 13 studies in HCWs from China, where more than one in every five healthcare workers suffered from anxiety or depression, with pooled prevalence rates of 23.2% for anxiety and 22.8% for depression [11]. Subsequent reviews reported broadly similar rates including a meta-analysis of 19 studies and estimated rates of 26% for anxiety and 25% for depression [53]. Even more surprising, however, was the finding that mental health concerns and anxiety symptoms in particular were more frequent in the Southeast Asian general population than HCWs. This pattern is at odds with previous observations elsewhere, whereby the rates were either similar or higher among HCWs compared to the general population during the same period of time. For example, Luo et al. [53] found that rates were akin between healthcare workers and the general public; though noted that studies from a number of countries such as China, Italy, Turkey, Spain and Iran reported higher-than-pooled prevalence among healthcare workers. Similarly, in their review, Vindegaard and Benros [57] concluded that HCWs generally appeared to experience more anxiety, depression, and sleep problems compared to the general population in a subgroup analysis of twenty studies. Furthermore, in our review, general and frontline staff recorded similar levels of psychological distress. Several previous studies demonstrated a higher psychological impact for frontline staff, yet others showed that the mental health effects of the crisis were equally felt across settings or specialties [58-60].

Overall, anxiety symptoms were more frequent than depression, a common finding across most studies to date [53]. Despite the considerable between-study heterogeneity, it appears that comparable proportions of respondents across groups recorded mild and moderate symptoms both for depression and anxiety, while more severe symptoms were less common.

Although insomnia was underreported in the studies under the scope of this systematic review, it was evidently the least prevalent mental disorder in Southeast Asia at 19%. Moreover, this rate compares favourably to the levels of 36% reported in the meta-analysis by Jahrami et al. [61] from 13 countries (Iraq, India, Germany, France, Italy, China, Mexico, Spain, Bahrain, Greece, USA, Australia, Canada) with further subgroup analysis highlighting the even greater frequency in Italy (55%) and France (51%). Overall, approximately two in five HCWs have been reported to experience some degree of sleep dysfunction [62], while shorter sleep duration has been associated with a higher likelihood of Covid-19 infection amongst HCWs [63].

### 4.3 Practical Implications

The results from this meta-analysis show that the rates of anxiety, depression and insomnia were lower in Southeast Asia compared to previous meta-analyses conducted in other areas. The disparity is particularly noticeable when compared to south European countries like Spain, France, Italy and Greece [64]. The differences between countries are likely multifactorial such as variation in pressures on healthcare systems, exposure to negative media and perceived lack of preparedness [65].

In addition, the lower prevalence rates in Southeast Asia could be associated to the recent experience with epidemics and the use of early interventions similar to those in China and east Asia. Indeed, some useful lessons could be learned from the interventions which were deployed throughout this region. Vietnam, for example, was lauded for its testing and surveillance system which was used to identify infection sources [66] and also recognised the importance of strengthening its grassroots healthcare system in order to contain Covid-19 [67]. In Singapore, the overall rates of preventative behaviours (e.g. avoiding public transport, social events and hospitals and reducing frequency/duration of shopping and eating out) were reportedly high [34], while another Singaporean study showed that the use of an official WhatsApp channel, providing information updates to the public, was protective against the development of depression [46]. According to Luo et al. [53] the use of precautionary measures to prevent the spread of Covid-19 and the access to up-to-date and accurate information were shown to shield from mental health problems.

Furthermore, a number of individual studies included in our systematic review, highlighted the mitigating role of higher levels of support against the development of anxiety symptoms. Amongst frontline HCWs, higher organisational and social support were both deemed to enhance resilience [27], and Sunjaya et al. [45] underlined the importance of general HCWs to maintain frequent contact with peers and families to prevent negative mental health effects. Social, family and governmental support were found to be protective in the student population [47], whilst living alone was a risk factor [36]. Furthermore, being single, separated or widowed was noted to be a risk factor within the general population [28] alongside increased exposure to Covid-19 information from the media which was associated with a higher likelihood of anxiety [25,30]. A separate study reported that a dose-response correlation was observed between information exposure of three or more hours per day and the severity of affective symptoms [43].

Finally, there is a number of Southeast Asian countries such as Myanmar, Cambodia, Laos, East Timor and Brunei without any available large-scale data on the mental health effects of the pandemic. For these countries without country-level studies, our systematic review on Southeast Asia may help them to use the results at the regional level as relevant evidence to guide their practice including the development of national mental healthcare strategies for pandemic-related interventions and short, medium and long-term service provision.

### 4.4 Strengths and Limitations

To our knowledge, this systematic review is the first to examine the pooled prevalence of depression, anxiety and insomnia in the general populations, HCWs and students during the COVID-19 outbreak in Southeast Asia. Despite the relative low number of studies per group and per country included in our meta-analysis, the total studies covered a considerable number of participants during a whole year of the pandemic. Furthermore, our subgroup analysis provided additional valuable insights of potential particular differences and /or vulnerabilities.

Nevertheless, there are some key limitations to our review. There was considerable disparity between the number of papers reporting on the four subgroups of populations, ranging from 14 (general HCWs), to 9 (general population), 6 (frontline HCWs) and only 3 (students). In addition, only two papers evaluated the presence of sleep problems, thus limiting the power of the findings on insomnia. Again, the majority of studies were conducted across inherently different countries at varying points in the course of the pandemic and some countries were not represented in this analysis which may limit the generalizability of our findings.

A variety of assessment tools were used to record the presence of mental health symptoms and different cut-off values were used to determine severity making it difficult to directly compare findings across studies. The quality of studies was also variable with high quality studies recording lower prevalence of mental health issues. Furthermore, non-English articles were excluded which could have created a bias. Finally, the studies included in our meta-analysis were all cross-sectional, thus the long-term physical and psychological implications of Covid-19 pandemic are not fully captured.

## 5. Conclusion

This systematic review is the first to report on the prevalence of anxiety, depression and insomnia in the general public and high-risk populations of Southeast Asia during the Covid-19 pandemic. The results of the meta-analysis demonstrate that a significant proportion experienced at least mild to moderate levels of anxiety and depression. However, the pooled prevalence revealed lower rates of mental health symptoms in the general population, healthcare workers and students in Southeast Asia compared to other areas such as China and Europe. Our findings can inform targeted identification of mental health symptoms and facilitate appropriate resource planning and allocation in the continued Covid-19 pandemic.

## Data Availability

All data are secondary and available per request.

## Credit author statement

SP: Writing – original draft, Writing – review & editing.

JC: Methodology, Validation, Formal analysis, Investigation, Data curation, Visualization, Writing – original draft, Writing – review & editing, Supervision.

JB: Writing – original draft, Writing – review & editing.

AD: Investigation (Data).

RKD: Investigation (Data).

WX: Investigation (Data).

AY: Investigation (Data).

BZC: Investigation (Data).

AD: Investigation (Data)

RZC: Investigation (Data).

SM: Investigation (Data).

XW: Investigation (Data).

SXZ: Conceptualization, Methodology, Validation, Formal analysis, Investigation, Data curation, Visualization, Writing – original draft, Writing – review & editing, Supervision.

All authors reviewed and approved the manuscript. The corresponding author attests that all listed authors meet authorship criteria and that no others meeting the criteria have been omitted.

## Transparency declaration

The corresponding author affirms that this manuscript is an honest, accurate, and transparent account of the study being reported; that no important aspects of the study have been omitted; and that any discrepancies from the study as planned (and, if relevant, registered) have been explained.

## Ethical approval

Not applicable

## Patient and public involvement

No patient or public was involved in a systematic review and meta-analysis

## Data and materials availability

All data are secondary and available per request.

**Table S1:** The search string used in this systematic review and meta-analysis. (Feb 6, 2021)

| Search | Search topic | Search keywords (titles, abstracts, and subject headings) with Boolean operators |
| --- | --- | --- |
| 1 | Exposure/<br>Context | 2019-nCoV OR 2019nCoV OR COVID-19 OR SARS-CoV-2 OR ((Wuhan AND coronavirus) |
| 2 | Outcome of<br>interest | "depressi*" OR "anxi*"OR "insomnia" OR "sleep symptoms" OR "sleep issue" OR "sleep problem" OR "mental symptoms" OR "mental issue" OR "mental problem" OR "psychiatric symptoms" OR "psychiatric issue" OR "psychiatric problem" |
| 3 | Epidemiological<br>phenomenon | “Prevalence” OR “Incidence” OR "rate*" OR "ratio*" OR “Epidemiolog*” OR “risk factor*” OR “relative risk” OR “odds ratio” OR “risk ratio” OR “disease burden” OR “Proportion” OR “percent*” |
| 4 | Language | English |
| 5 | Time | 2020.2.1 – 2021.2.6 |
|  | Overall search | 1 AND 2 AND 3 AND 4 AND 5 |

